# Early Prediction of Anti-PD-1 Response in Hepatocellular Carcinoma via Multi-Rank Taxonomic Feature Engineering and a LUT-Based Prediction Vehicle

**DOI:** 10.1101/2025.05.15.25327728

**Authors:** Phing Chan Chai, Oleg V. Moskvin, Rohan Williams, Damien Keogh

## Abstract

**Background:** Hepatocellular carcinoma (HCC) is a leading cause of cancer-related mortality. Anti-PD-1 therapy is a standard option for advanced HCC, but response rates are modest and pre-treatment response prediction remains an unmet clinical need. Prior gut microbiome-based models have failed to discriminate responders from non-responders at baseline, limiting their clinical utility for early treatment planning.

**Methods:** We reanalyzed 16S rRNA sequencing data from 35 HCC patients receiving anti-PD-1 therapy (155 longitudinal samples [1]). Key methodological choices to enable model performance and results reliability: (1) multi-rank taxonomic features to address taxonomic assignment uncertainty and rectify informative signal by combining taxonomic resolutions, (2) log-transformation to leverage low-abundance but informative taxa routinely lost by conventional pipelines, and (3) strict subject-level train-test partitioning to prevent longitudinal-sample leakage. To enable standalone deployment, we distilled the trained neural network into a compact lookup table (LUT) with frozen Centered Log-Ratio normalization and N-linear interpolation.

**Results:** A neural network classifier built on 8 selected taxa achieved AUROC 0.784 ± 0.024 on held-out test data. Critically, the model discriminated responders from non-responders at baseline (AUROC 0.681 ± 0.056), a qualitative shift from the original analysis (∼0.5 at baseline). Per-patient predictions were stable across treatment timepoints, indicating the signature reflects intrinsic microbiome composition rather than drug-induced perturbation. The 2,916-row LUT reproduced the neural network’s discrimination in an end-to-end pipeline from raw abundance counts to response probabilities.

**Conclusions:** This work delivers two contributions with distinct validation requirements. The 8-taxon signature demonstrated for the first time that taxonomy cross-rank pooling can yield a measurable response signal before the intervention start. This qualitative result is primed for prospective external validation and quantitative improvement. Independently, the neural-network-to-LUT distillation, with frozen Centered Log-Ratio normalization and N-linear interpolation, is a standalone computational contribution independent of any signature it compresses, providing a transparent and explainable deployment vehicle for neural classifiers.

**Trial registration:** Not applicable. This study is a secondary analysis of publicly deposited 16S rRNA gene sequencing data from a previously reported prospective cohort [1]; no clinical intervention was performed by the present authors.

## Introduction

Hepatocellular carcinoma (HCC) is a major global health concern, being the sixth most common cancer and the third leading cause of cancer-related deaths worldwide. The prognosis for advanced HCC remains poor, with limited effective treatment options historically available[2]. Immune checkpoint inhibitor (ICI) therapy has reshaped the systemic management of advanced HCC. Beyond early monotherapies targeting programmed cell death protein 1 (PD-1) and its ligand (PD-L1), combination regimens, e.g. atezolizumab plus bevacizumab and tremelimumab plus durvalumab, are now first-line standard of care, although sustainable benefit is still confined to a minority of patients [3]. ICIs, such as those targeting programmed cell death protein 1 (PD-1) and its ligand (PD-L1), have shown significant potential in reactivating the immune system to target and destroy cancer cells [4].

Despite the promise of ICIs, a significant proportion of patients with HCC do not respond to these therapies, exhibiting either primary or acquired resistance [5]. This resistance poses a major challenge as it limits the overall efficacy of ICIs and complicates treatment planning. Identifying patients who are likely to be resistant to ICIs before starting therapy is crucial for optimizing treatment strategies and improving outcomes [6].

Currently, host-derived biomarkers, including genomic variants, expressed proteins, and enrichment of specific host cell types in the tumor microenvironment, have been utilized to predict responses to ICIs. Examples include tumor mutational burden (TMB), microsatellite instability, and genetic variants in several specific pathways [7], the expression level of PD-L1 itself [8], and the abundance of CD8+ T cells [9]. However, these biomarkers often have limited predictive power or require invasive tissue biopsies. The role of microbiota in response to anti-PD-1 therapy was noted as early as 2018, when higher abundances of *Fecalibacterium* and *Bacteroidales* were associated with longer progression-free survival [10]. Other studies have demonstrated that the diversity and specific composition of the gut microbiota can significantly affect the effectiveness of ICIs by modulating host immune responses. Notably, enrichment of specific commensals, such as *Faecalibacterium* and *Akkermansia muciniphila*, as well as overall microbial diversity, has been correlated with an enhanced response to PD-1 blockade across several cancers [11], [12]. Currently, the gut microbiome is being considered as a potential source of biomarkers for predicting responses to immune checkpoint inhibitors [13]. The biological mechanisms of microbiota-host interactions that affect immunotherapy responses are reviewed in recent publications [14] [15].

Moreover, stool sample collection and analysis to assess the gut microbiota offer several practical benefits. Unlike tissue biopsies required for genomic biomarker analysis, stool sample collection is a non-invasive, low-cost, and readily available method. Thus, the inclusion of microbiome-based assays for response prediction could add significant value to clinical practice.

Research has identified microbiome-based therapy response predictors for various cancers, underscoring the potential of this approach in oncology. For instance, a network-based microbiome-centric survival predictor outperformed traditional host-related factors such as PD-L1 levels, antibiotic use, and age in predicting outcomes in non-small cell lung cancer [16]. Similarly, in melanoma, metagenomic studies have highlighted the role of specific bacterial taxa in influencing therapy response [17]. Importantly, existing research highlights causal links between the gut microbiota and cancer immunotherapy, beyond mere statistical associations, as demonstrated by Mendelian randomization studies [18] and improvement of therapy response after direct bacterial supplementation [19].

However, a recurring limitation of microbiome-based ICI predictors has been their poor cross-cohort generalizability, reflecting a combination of biological inter-cohort heterogeneity, technical processing biases, and methodological pitfalls, including inadequate train/test partitioning [20], [21]. Recent consensus reviews have identified rigorous feature engineering, leak-free validation, and deployment-ready models as the field’s central translational bottlenecks [22], [23]. In hepatocellular carcinoma, microbiome-based predictors are still in the research stage, but early findings are promising. While studies have implicated the gut microbiota in ICI response in hepatobiliary cancers, including reports of *Bacteroides stercoris* and *Parabacteroides merdae* enrichment in atezolizumab/bevacizumab responders [24], the development of robust, deployable predictive models remains ongoing [3].

The primary reason for the absence of a clinically ready model is the poor performance of reported models at baseline. In this study, we developed a predictive framework using publicly available microbiome and clinical data from patients with HCC undergoing anti-PD-1-based immunotherapy [1]. Our approach, based on custom feature engineering and machine learning methodology, demonstrates the feasibility of a robust model that predicts response to anti-PD-1 therapy from samples obtained before the onset of drug intervention, a finding not achieved in the original analysis. Furthermore, we distilled the trained model into a portable lookup table (LUT) that enables standalone deployment from raw microbiome counts to therapy response probabilities without requiring the original neural network infrastructure. Thus, we directly address the need for deployment-ready, interpretable microbiome diagnostics highlighted in recent clinical-translation roadmaps [22].

We addressed the known outstanding challenges of predicting anti-PD-1 therapy response from gut microbiome data that impede clinical applications: (a) the granularity of microbial features that are sparsely distributed between individuals, (b) the large number of features in existing models, (c) the absence of a predictive signal at baseline, and (d) the lack of a deployment-ready prediction framework. This was achieved by (a) considering a set of features across taxonomy ranks, (b) incorporating features with lower abundance but high prevalence via data transformation, (c) designing a custom machine learning architecture, and (d) constructing a LUT-based standalone prediction engine with frozen normalization parameters. Our analysis provides a substantive advance over the findings of the source study, which detected a response-related signature only in subjects after the commencement of drug treatment.

## Methods

### Dataset

We used publicly available data from Wu et al. [1], a study of 35 patients (31 males, 4 females; mean age, 53.8 years) with hepatocellular carcinoma (HCC) who underwent anti-PD-1-based immunotherapy. The majority of patients had advanced-stage HCC, classified as Barcelona Clinic Liver Cancer stage C (32/35, 91.43%).

The distinction between responders (R, 46% of the cohort) and non-responders (NR, 54%) was performed in the original publication based on a combination of complete response (CR), partial response (PR), or stable disease (SD) labels and follow-up time [1]. The 46%/54% distribution indicated a relatively balanced dataset, enabling robust comparative analyses. Notably, the R and NR groups were also balanced by age, sex, liver function status, and underlying etiologies, except for baseline alpha-fetoprotein (AFP) levels, immature granulocyte counts, and lymphocyte counts.

The raw 16S rRNA gene sequencing data representing this study in the National Genomics Data Center (NGDC) Genome Sequence Archive (GSA) database (accession: CRA005195) is accompanied by a limited metadata file. By applying the knowledge of the three-week interval between fecal sample collections, we were able to match the samples to all 35 patients (Supplemental Data 1). The number of anti-PD-1 injections and corresponding fecal samples varied among patients, as illustrated in Supplementary Figure 1.

### Microbiome data analysis

Raw sequencing data (FASTQ files) were downloaded from GSA and reprocessed using the BluMaiden standard 16S amplicon analysis workflow. Low-level data preprocessing followed the BluMaiden KEYSTONE™ pipeline. Modifications aimed at optimizing the predictive model’s performance are described in the Results section.

Data analysis was performed using a combination of the R statistical environment [25] and the Python programming language, with custom scripting and open-source packages. Community diversity measures and PERMANOVA analyses were calculated and performed using the Scikit-bio package (ver. 0.6.0).

The differential abundance of microbiota features relevant to clinical phenotypes was assessed in a multivariable analysis setting using MaAsLin2 [26] version 1.14.1. The analysis was run with *min_abundance* = 0 and *min_prevalence* = 0.1 parameters, setting the factors of interest (Responder Status and Doses Taken) as target variables. Because a custom normalization approach was used, internal standardization was disabled by setting *standardize = FALSE*. Patient ID was included as a random effect.

### Predictive modeling

Custom ML model construction was performed using Python programming language, open-source packages, and in-house developments of BluMaiden Biosciences. The neural network architecture consisted of fully connected layers with ReLU activation functions, trained using the Adam optimizer with early stopping based on validation loss. Hyperparameter optimization was performed via grid search over learning rate, layer sizes, and dropout rates. Feature importance was assessed using Shapley Additive exPlanations (SHAP) values. The model was evaluated on the test dataset using AUROC scores to measure predictive accuracy. Unless stated otherwise, the AUROC scores are represented as mean +/- standard deviation across 100 independently trained models, each fitted on different train/validation resampling drawn from the fixed training set and evaluated on the same held-out test set. The dispersion, therefore, reflects variability across training subset resampling and training stochasticity. To visualize the prediction output for individual samples, a binary classification of responder status was generated based on a probability score (below or above 0.5) mapped via a sigmoid function. Biomarkers identified through predictive modeling have been claimed in United States Patent Application No. 63/762204.

### Lookup table construction and deployment

To translate the neural network predictor into a standalone, model-agnostic prediction tool, we distilled the trained model into a compact lookup table (LUT). The LUT was constructed by identifying informative regions in the normalized abundance values for each of the eight features through frugal adaptive quantization, which preserves feature-level prediction informativeness while minimizing table size. The resulting 2,916-row LUT (Supplementary Table 1) encodes the key combinations of discretized input values and their corresponding predicted therapy response probabilities.

Normalization of input data for LUT-based prediction follows a frozen pipeline that exactly reproduces the training-time procedure, ensuring transferability to arbitrary new datasets:

(1) Centered Log-Ratio (CLR) transform — computed over all taxa in the sample (not just the 8 model features):

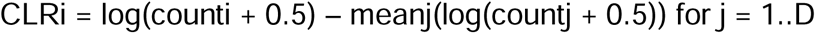

where D is the total number of taxa columns in the input.

(2) Feature selection — the 8 model features are extracted from the full CLR matrix.
(3) Min-max scaling — each of the 8 CLR values is scaled to [0, 1] using frozen min/max constants derived from the discovery cohort:

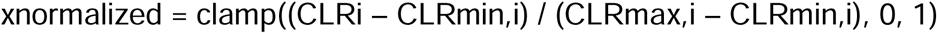

The frozen CLR boundaries for each feature are reported in Table 1.

**Table 1.** Frozen CLR boundaries for min-max normalization of the 8 model features, derived from the discovery cohort (n = 155).

| Feature | CLR min | CLR max |
| --- | --- | --- |
| *Acidaminococcaceae* | -1.6448 | 6.1041 |
| *Ligilactobacillus* | -1.8309 | 8.0387 |
| *Pasteurellaceae* | -1.8309 | 7.4395 |
| *Dorea* | -1.7690 | 5.9186 |
| *Barnesiella* | -1.8309 | 4.5234 |
| *Tannerellaceae* | -1.6913 | 7.0897 |
| *Ruminococcus* | -1.7690 | 7.0478 |
| *Atopobium* | -1.9479 | 2.7532 |

Interpolation between LUT vertices is performed in unbounded space rather than directly in the bounded probability space, to improve interpolation accuracy in the inherently non-linear probability domain [27]. Specifically, the stored (strictly positive) outcome probabilities are mapped to natural log-probability space prior to N-linear interpolation; the interpolated vector is mapped back to a normalized probability distribution via softmax [28].

## Results and Discussion

### Microbiome compositional differences in clinical groups

Conventional characterization of gut microbiome compositional differences between clinical groups involves both ecological measures (species diversity, richness, and evenness) and associations of abundance levels of microbial features with clinical parameters via analysis of variance and mixed linear models. The original study applied these methods and confirmed their inadequacy for deriving clinically applicable biomarkers, whereas nonlinear machine learning methods demonstrated practical potential. Since our findings confirm this observation, we briefly discuss traditional descriptive approaches to (a) establish a common baseline with the published study, (b) explore deeper taxonomy representation that includes multiple taxonomy ranks to balance generalizability and precision, and (c) leverage the updated status of taxonomy interpretation of the same sequencing data.

Figure 1A shows the results of ecological parameter analysis, which included Shannon diversity, Chao1 species richness, and Simpson’s evenness. Responders and non-responders exhibited similar levels of baseline microbial diversity at the genus level. While the Shannon Diversity index indicated that responders had slightly higher diversity, none of the differences in all ecological measures between responders and non-responders were statistically significant (Mann-Whitney U test: Shannon, *p*=0.253; Chao1, *p*=0.619; Simpson’s, *p*=0.456), which is consistent with the findings of the original study performed at the OTU level. The non-informativeness of overall ecological measures was further supported by the lack of differences in Shannon diversity between the two groups after examination that included: (a) consideration of five taxonomic ranks (from phylum to genus) and (b) separate assessment of both the entire cohort and a subset of baseline samples before receiving the first dose (Supplementary Figure 2).

**Figure 1:**
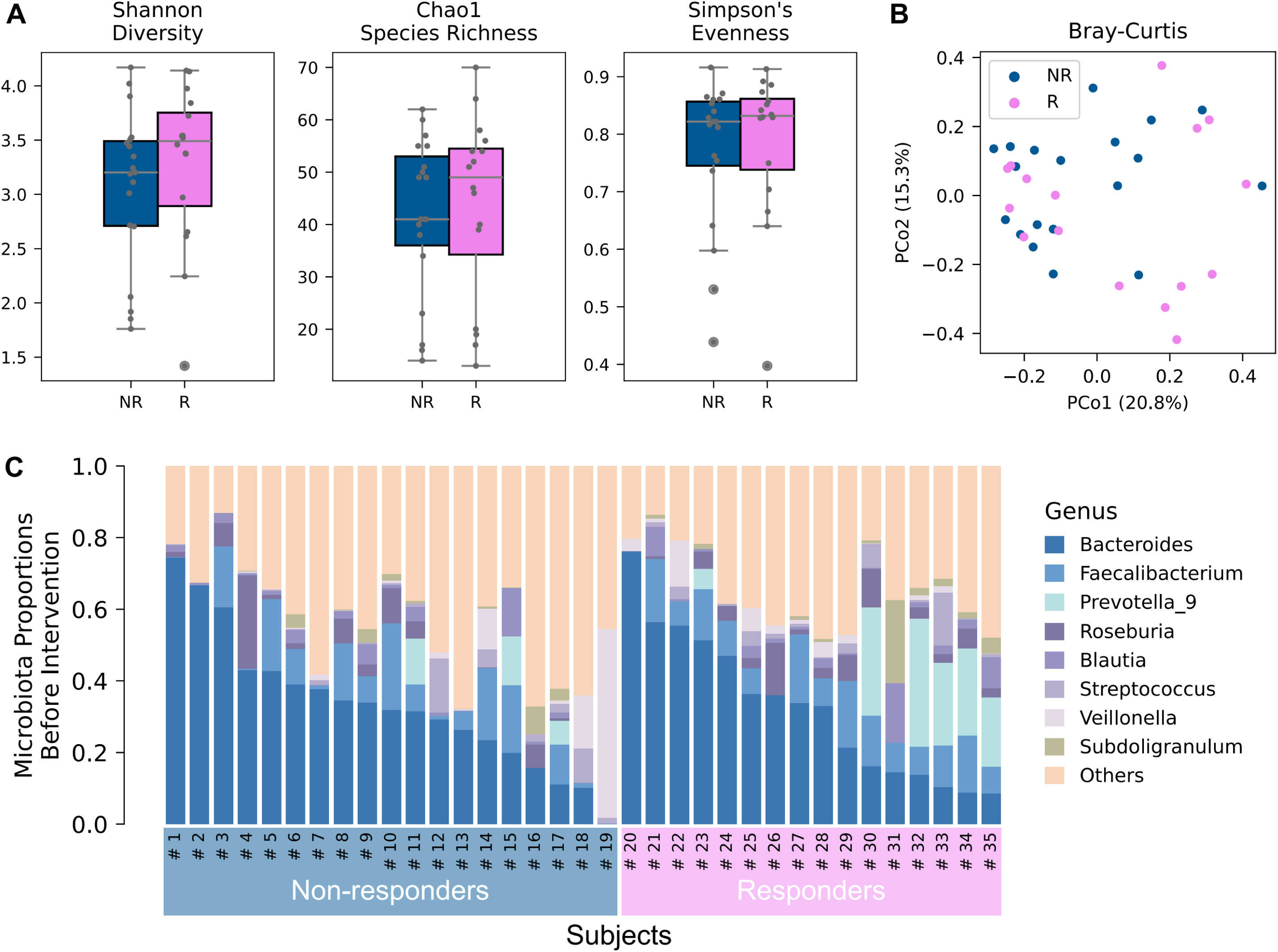
Baseline microbial diversity and composition analysis. (A) Alpha diversity metrics comparing responders (R) and non-responders (NR) at baseline. Boxplots display Shannon Diversity, Chao1 Species Richness, and Simpson’s Evenness. (B) Principal Coordinate Analysis (PCoA) plot of microbial community composition at baseline using Bray-Curtis dissimilarity. Each dot represents a sample (blue: NR; pink: R). (C) Stacked bar plot showing proportions of microbial genera in each patient at baseline. Top eight genera are displayed, with all others aggregated as ‘Others’.

Beta diversity was assessed to probe differences in microbial community composition between responders and non-responders. The principal coordinate analysis (PCoA) plot based on Bray-Curtis dissimilarity (Figure 1B) revealed no significant differences in microbial communities between the two groups at baseline (PERMANOVA: F = 0.988, p = 0.391). Other dissimilarity metrics, including Jaccard distance and weighted UniFrac, were also not helpful in revealing differences at baseline (Supplementary Figure 3). Additionally, compositional representations of neither the top genera (Figure 1C) nor other taxonomic ranks (Supplementary Figure 4) revealed microbiome compositional patterns associated with therapy response. Taken together, both alpha and beta diversity measures at baseline failed to significantly stratify patients into responders and non-responders.

Similar to observations in the original publication on accumulation of microbiome differences between R and NR groups following the course of intervention, we also detected an association between responder status and microbiome composition of drug-exposed individuals. Significant differences in microbial community composition between R and NR groups were detected in the entire pool of samples using either Bray-Curtis dissimilarity (PERMANOVA: F=2.793, p=0.004) or weighted UniFrac distance (PERMANOVA: F=4.251, p=0.004). These results indicated that microbial communities of responders and non-responders diverged more noticeably when the entire course of treatment was considered (Supplementary Figure 3).

In addition to group-level differences, we examined whether associating individual microbial features with therapy response could reveal a statistically significant signal. For this purpose, we applied generalized linear modeling implemented in the MaAsLin2 package. This analysis revealed that none of the genera were significantly associated with either responder status or the number of anti-PD-1 doses taken (Figure 2). The absence of significant associations in this multivariate analysis (q: 0.428–0.998 for responder status; 0.456–0.962 for doses taken) adds another level of microbiome compositional representation that, similar to more generalized parameters, is insufficient to detect a signal of response. This highlights the complexity of the fecal microbiome and its interaction with clinical outcomes, suggesting that more sophisticated models may be required to uncover meaningful biomarkers for patient stratification.

**Figure 2:**
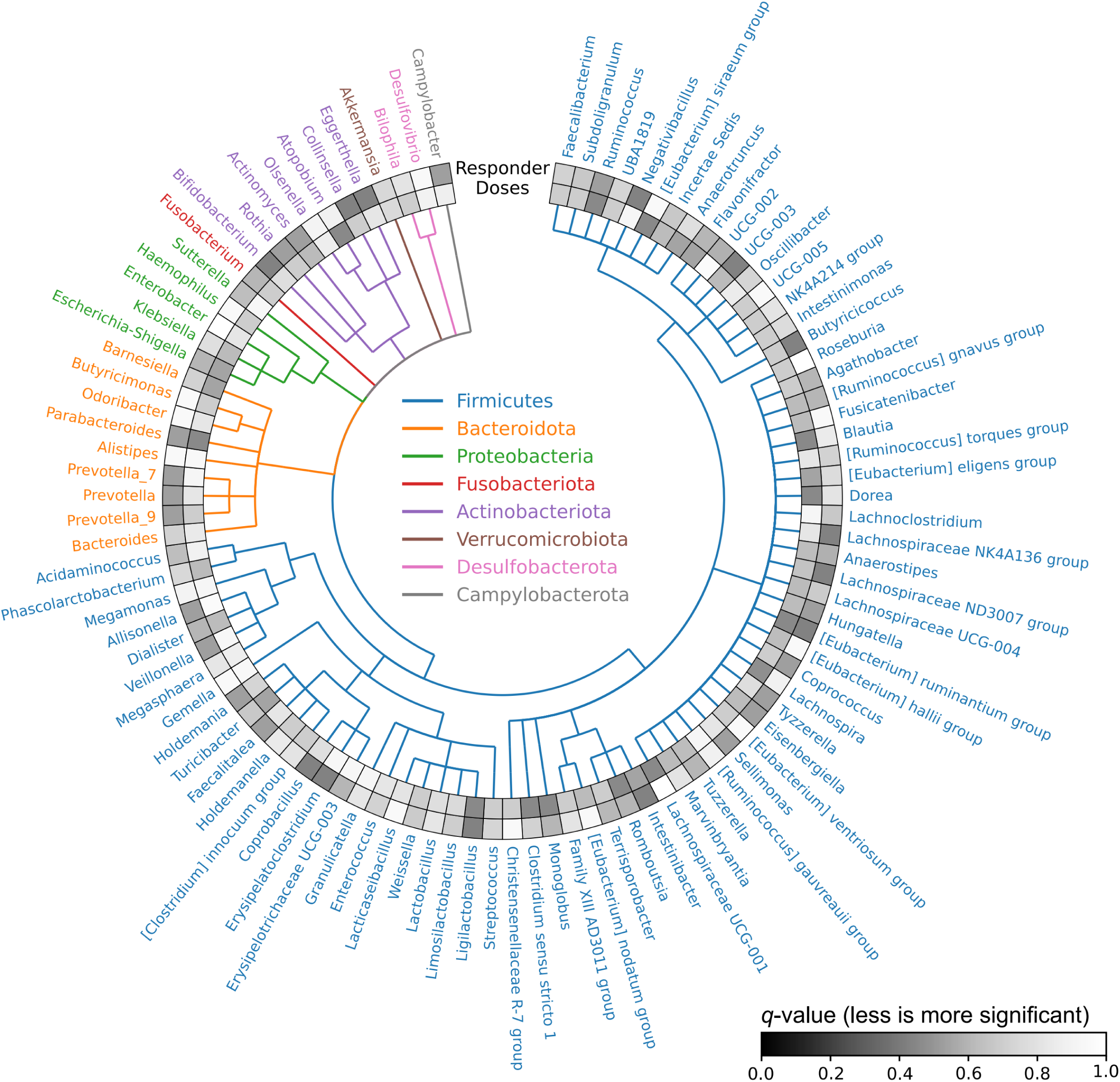
Phylogenetic tree with multivariate analysis heatmap. The phylogenetic tree displays 100 genera filtered to remove low-abundance taxa, with branches colored by phyla. The outer heatmap shows q-values from MaAsLin2 analysis assessing associations between Responder status, anti-PD-1 Doses, and genus abundance (darker shades represent lower q-values, more significant).

### Microbiome-based predictor of treatment response

The set of study subjects had the favorable characteristic of being balanced across most clinical parameters. This balance establishes the stage for discovering robust clinical biomarkers of therapeutic response that are not biased by other clinical differences. If the challenge of the association signal’s invisibility can be successfully resolved using less conventional approaches, the resulting markers are expected to be unbiased and applicable for patient stratification.

To reveal the clinically applicable signal, we addressed the following challenges: (1) conservative partitioning of subjects into training and test sets while preventing any data leakage, (2) overcoming the high granularity of taxonomic features when observed across subjects, (3) finding the right balance between features’ within-sample abundance and across-sample frequency of detection, and (4) optimizing the learning architecture.

### Partitioning samples into train and test sets

To build an ML-based predictive model, partitioning patients into suitable training and test sets was critical given the high variance in the number of anti-PD-1 doses administered to each patient. In the original article by Wu et al., the authors did not specify how they conducted their train-test data split beyond mentioning a 70:30 train-validation split, presumably performed at the sample level randomly. This approach poses a risk of optimistic bias, as samples from the same patients at different time points could end up in both training and validation sets. Since fecal microbiome profiles tend to be more invariant within subjects than between subjects (PERMANOVA, p = 0.001), it is essential to assign all samples from a single patient exclusively to either the training or the test set.

To adequately represent both responders and non-responders in training and test sets while preventing data leakage, we used heuristic optimization to split samples into 109 samples from 24 patients in the training set and 46 samples from a distinct group of 11 patients in the test set. The resulting 70:30 train-test split demonstrated balanced distribution across both response groups, overall number of samples, and other metrics (Figure 3A-B). This robust train-test split mitigates risk of data leakage and maintains balanced representation, thereby ensuring real-world applicability of the generated model.

**Figure 3:**
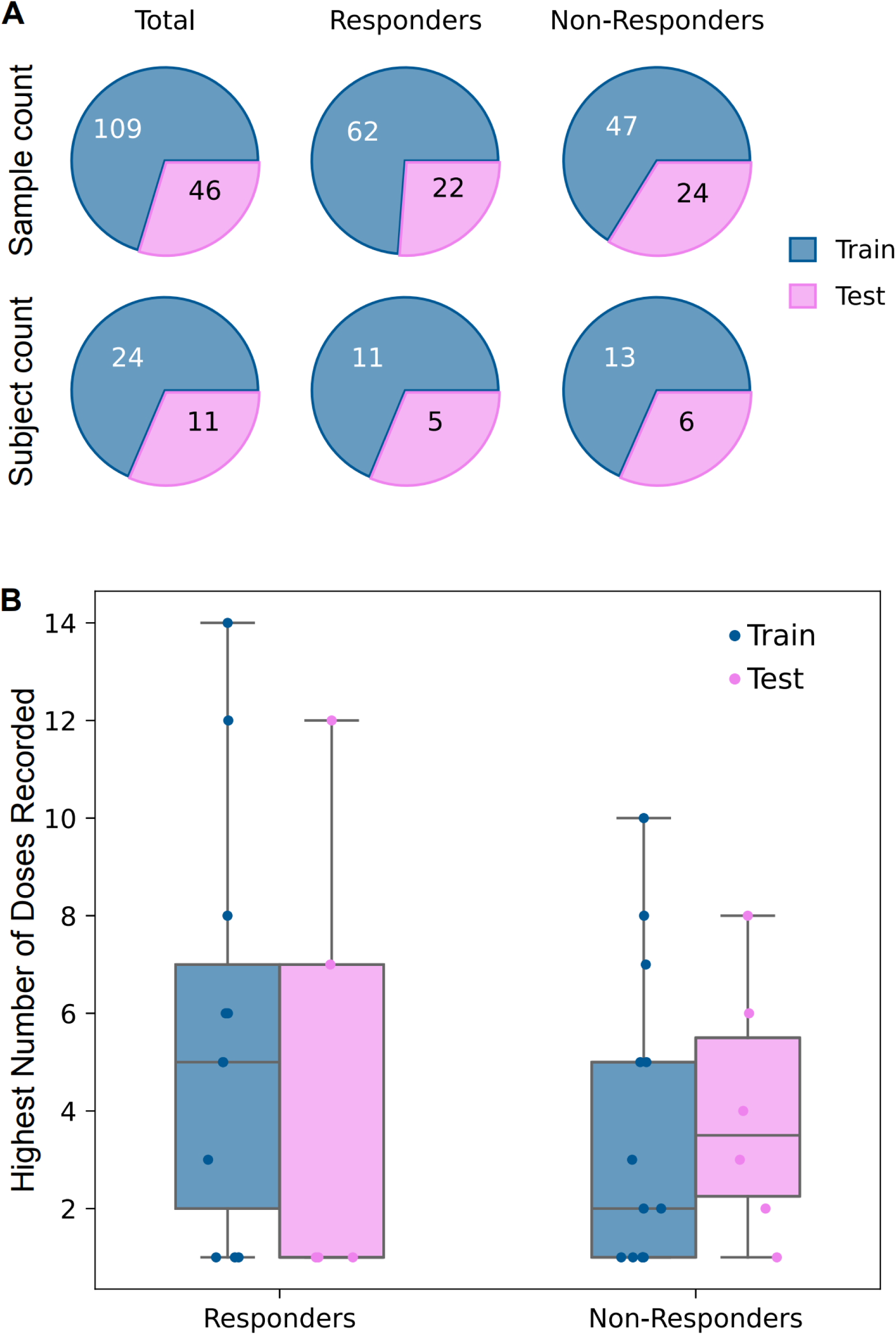
Sample and subject distribution in train-test split. (A) Pie charts showing the distribution of samples and subjects in the train and test sets for the total cohort, responders (R), and non-responders (NR). (B) Box plots showing highest number of anti-PD-1 doses recorded for R and NR in both train (blue) and test (pink) sets. The distribution illustrates the balance achieved in the train-test split, with both sets maintaining similar dose distribution profiles for R and NR.

We leveraged various established auto-ML libraries as benchmarks to establish a realistic baseline for model performance. AutoGluon, TPOT, auto-sklearn, and mljar-supervised produced best models with AUROCs ranging from 0.479 to 0.696 on our leak-free test set of 408 features (Figure 4A). The fact that auto-ML AUROCs could be close to 1.0 on the validation set but collapse on the test set clearly demonstrates the overfitting challenge that out-of-the-box methods cannot properly address. Improving the poor performance of conventional methods required custom decisions regarding both the model architecture and the microbiome data representation.

**Figure 4:**
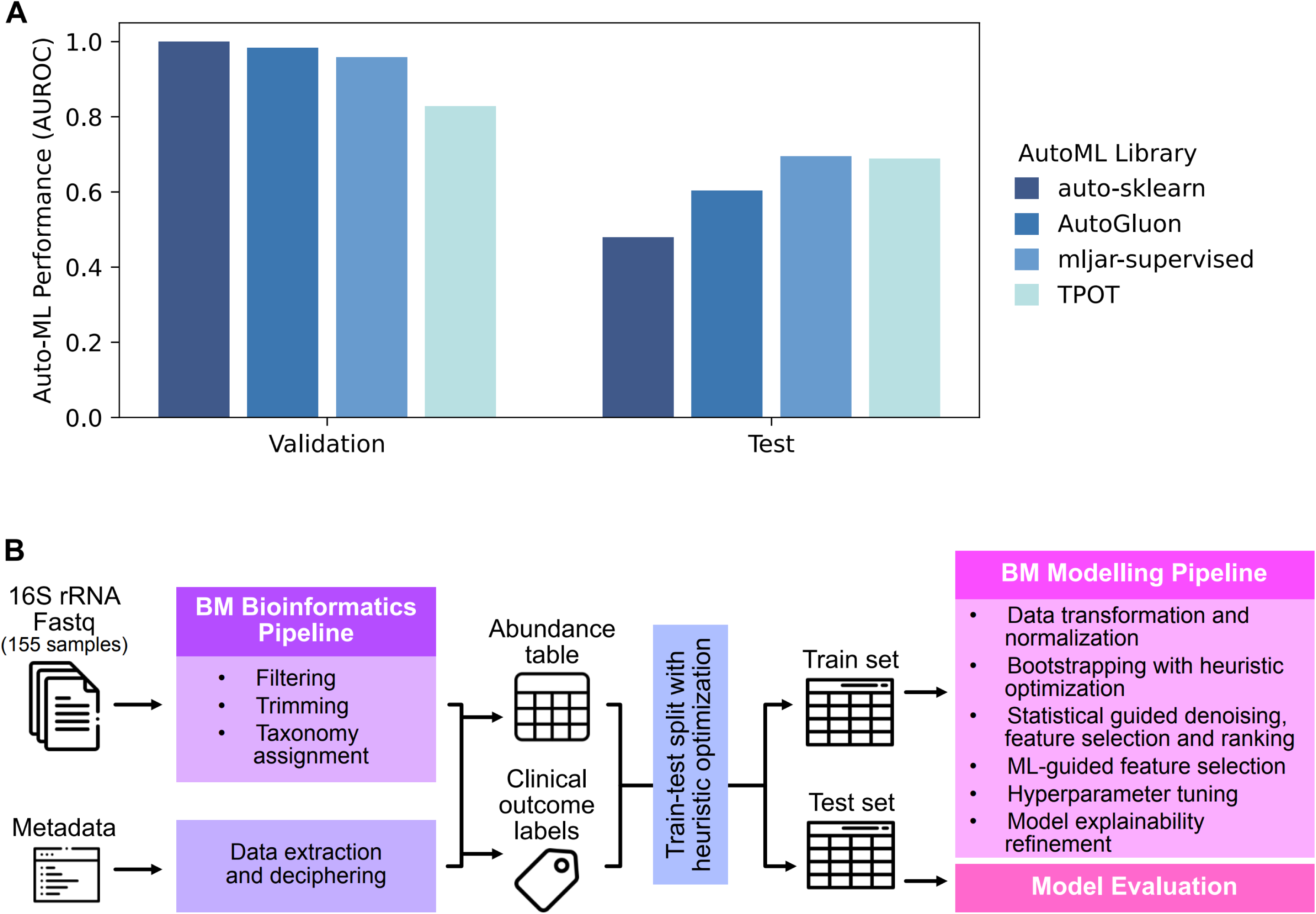
AutoML benchmark performance and modeling workflow. (A) Bar plot showing AUROC performance of best models from four AutoML libraries (auto-sklearn, AutoGluon, mljar-supervised, TPOT). (B) Conceptual overview of BluMaiden pipelines for 16S rRNA data processing and modeling.

### Predictive modeling

To develop robust predictive models, we carefully processed microbiome abundance data to extract meaningful features while addressing common challenges in fecal microbiome analysis. First, we used microbial taxa from phylum to genus ranks as features. Although this method introduces overlapping features, it is necessary because of inherent uncertainty in taxonomic assignment and partial mutual support of less informative (but more stable) features at higher taxonomic ranks, as well as potentially more informative (but sparse across individuals) features at lower taxonomic ranks. Moreover, due to resolution limitations of 16S metabarcoding data, sequencing reads often remain unclassified at lower taxonomic ranks while being represented by aggregated counts at higher ranks. By incorporating features from multiple taxonomic levels, we captured a more complete picture of microbial community structure with reduced sparsity. This approach resulted in a pool of 408 taxonomic features: 14 phyla, 21 classes, 55 orders, 92 families, and 226 genera (Figure 4B).

Because of the notorious interpersonal variability of microbiome profiles, each individual in the training set had, on average, only 25.5 ± 8.0% of total features represented. This sparsity is a common issue in microbiome data and can lead to significant data loss if not properly managed. A common practice of filtering out low-abundance features risks discarding taxa that may still provide valuable information. To retain the latter, we applied sample-wise log transformation to stabilize variance, normalize the distribution of feature counts, mitigate the influence of extreme values, and ensure retention of low-abundance but potentially informative taxa (Figure 4B).

Despite these preprocessing steps, many microbial features may still be affected by noise from sequencing or uncertain taxonomic assignments. To address this, we employed our suite of statistical and machine learning tools to identify and rank features based on their association with and informativeness of the predictive outcome. Together, these techniques reduced the number of features from 408 to 8, which constituted the core microbial taxa that could serve as biomarkers for detecting responders versus non-responders to anti-PD-1 therapy among HCC patients.

Our final model was a neural network utilizing these eight key taxa as features. After hyperparameter tuning, the performance of this model, as illustrated in the ROC curve (Figure 5A), achieved an AUROC of 0.784 ± 0.024 for the test data. This result indicates good predictive capability, significantly outperforming all auto-ML benchmarks (auto-sklearn, 0.479; AutoGluon, 0.604; mljar-supervised, 0.696; TPOT, 0.689). Our test AUROC of 0.784 reflects performance under strict subject-level separation, which provides a conservative but realistic estimate of generalization performance. In contrast to Wu et al.’s model utilizing 18 biomarkers, our model’s use of only eight taxa (across taxonomic ranks) offers several practical benefits. Our simpler model, which includes fewer than half the biomarkers, reduces complexity and cost of clinical testing while enhancing interpretability.

**Figure 5:**
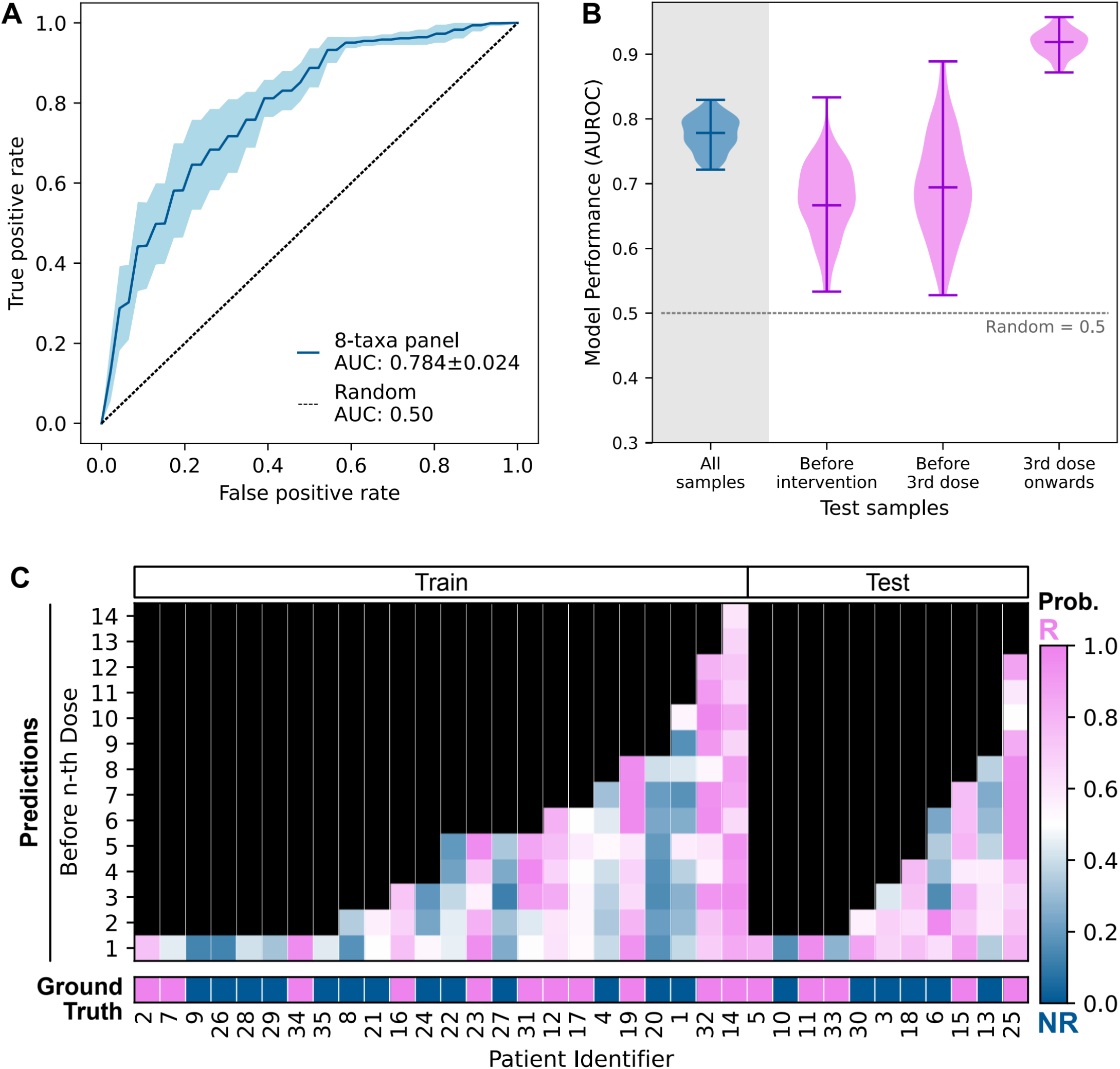
Neural network model performance and predictions. (A) ROC curve showing performance of 8-taxa neural network model (AUROC 0.784 ± 0.024). Blue line: mean performance across 100 trials; cyan shading: standard deviation. (B) Violin plots of AUROC scores across 100 models for different sample groups (all samples, baseline, before 3rd dose, 3rd dose onwards). (C) Heatmap of predicted R/NR status with probability threshold 0.5. The probabilities are the mean of 100 trials. Bottom bar shows ground truth responder status (pink: R; blue: NR).

The baseline AUROC of 0.681 ± 0.056, while modest in absolute terms, represents a meaningful advance for non-invasive pre-treatment stratification. For comparison, PD-L1 expression — the most widely used ICI response biomarker — achieves comparable or lower discrimination in many tumor types and requires invasive biopsy. The clinical value of our microbiome signature lies not in replacing established prognostic factors but in providing an orthogonal, non-invasive information source that could be integrated into multimodal prediction frameworks.

The qualitative advantage of our model over that of the original publication is its ability to detect the response signal before starting drug intervention. The model by Wu et al. was unable to detect this signal (random-chance AUROC ≈ 0.5). The ability to detect therapy response signal in microbiome data at baseline provides a tremendous advantage to the clinical utility of such models, especially when combined with parsimonious feature composition, enabling simpler and cheaper implementation of the real-world test.

### Insights into informative taxa and clinical utility projection

Our eight biomarkers included three taxonomic features at the family level (*Acidaminococcaceae*, *Pasteurellaceae*, and *Tannerellaceae*) and five features at the genus level (*Ligilactobacillus*, *Dorea*, *Barnesiella*, *Ruminococcus*, and *Atopobium*). In contrast, the model of Wu et al. exclusively utilizes biomarkers at the genus level.

All three of our family-level biomarkers had corresponding genus-level biomarkers (one for each family) used in Wu et al.’s model (Figure 6A). These family-level markers have potential to capture a broader range of bacterial species with similar biological properties and predictive values, thereby overcoming the granularity of genus-level features across individuals. Notably, the most informative marker based on SHAP values in our study, *Acidaminococcaceae*, maps to two taxa at the genus level, *Acidaminococcus* and *Phascolarctobacterium*, with only the latter identified as a key microbial biomarker (though the least important) in Wu et al.’s study. This indicates that our pan-taxonomic-rank approach can leverage features that capture broader microbial diversity.

**Figure 6:**
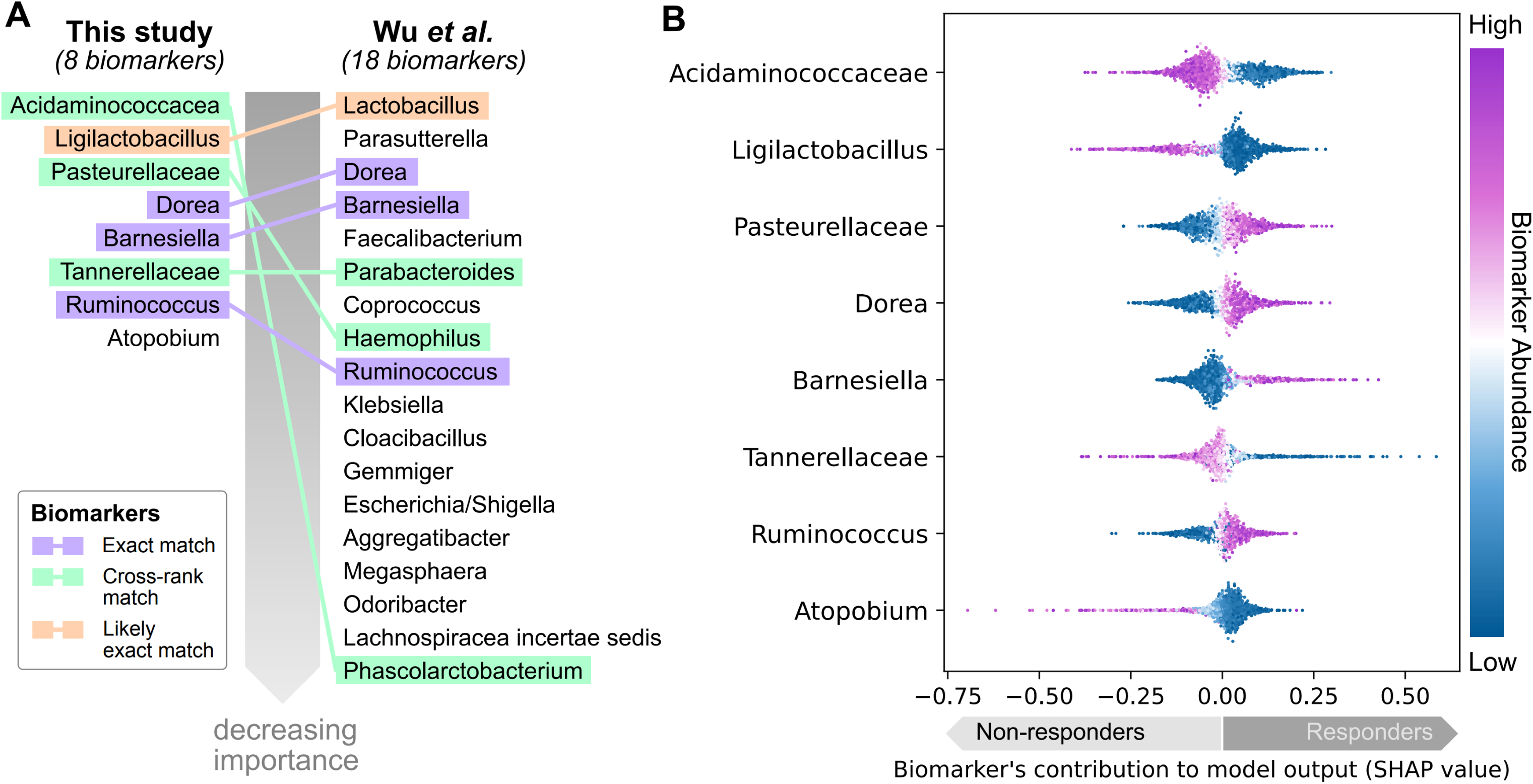
Biomarker importance from SHAP analysis. (A) Comparison of 8 biomarkers identified in this study with 18 biomarkers from Wu et al., arranged by decreasing importance. Colored lines indicate exact matches, cross-rank matches, and high phylogenetic closeness. (B) SHAP beeswarm plot showing impact of 8 biomarkers on model output. Color indicates biomarker abundance (purple: high; blue: low). Positive SHAP values indicate a contribution toward responder prediction. The points are an aggregation of predictions from the validation samples when the model was trained on a heuristic-optimized bootstrapped training dataset.

For our genus-level biomarkers, three (*Dorea*, *Barnesiella*, and *Ruminococcus*) overlapped with key microbial biomarkers of Wu et al., highlighting their significance and providing convergence for identification of crucial biomarkers. Interestingly, our method identified *Ligilactobacillus* as the second-most informative biomarker, whereas Wu et al.’s model identified *Lactobacillus* as the most important biomarker. *Ligilactobacillus* is taxonomically close to *Lactobacillus* and has historically been reclassified from the latter. Similar to *Lactobacillus*, it has a substantial record of probiotic properties [29]. Taxonomic closeness and the respective functional overlap between the two genera suggest that both models may capture the same underlying biological signal, albeit with significantly different practical applicability.

The SHAP value plot provides insights into the contribution and impact of each biomarker on model output (Figure 6B). The top contributors to the model’s predictions, *Acidaminococcaceae* and *Ligilactobacillus*, were negatively associated with responder status. *Tannerellaceae* and *Atopobium* also showed negative associations with responders. Conversely, *Pasteurellaceae*, *Dorea*, *Barnesiella*, and *Ruminococcus* positively contributed to the model-predicted responder status.

The detection of *Acidaminococcaceae* as a family-level biomarker of non-response makes sense given that one genus of this family, *Phascolarctobacterium*, was captured in the same context by Wu et al., while another genus, *Acidaminococcus*, was reported to be associated with adverse effects of immune checkpoint inhibitor therapy, being enriched nearly 200 times in the Adverse Events group [30]. Despite this functional connection, we prefer to avoid over-analyzing the composition of our biomarker signature based on existing literature. Given the nature of the neural network model, which relies heavily on complex non-linear relationships between marker feature abundance and clinical phenotypes, the involvement of network-level relationships is reasonably expected. This makes functional interpretation of the signature taxa a longer path that requires specific, focused studies.

Among the eight biomarkers, detection of individual markers showed a median prevalence of 67.8% across all patients. In comparison, the median prevalence of biomarkers from Wu et al.’s model was substantially lower at 51%, with some markers (such as *Aggregatibacter* and *Parasutterella*) dropping to be detectable in only 1.94% of patients — over 6 times less than the minimal prevalence observed for our model’s markers (12.3%). This sparseness of markers in the original model is expected to decrease the consistency of predictive power across different patient cohorts.

### Temporal stability of predictions and individualized microbiome signatures

The robustness of our model was additionally confirmed by its performance across different stages of anti-PD-1 intervention, which showed useful degree of predictive power even at early stages (Figure 5B), such as baseline (AUROC: 0.681 ± 0.056) and before the third dose of anti-PD-1 (AUROC: 0.688 ± 0.058). This qualitatively differs from the original model, which performed randomly with AUROC approximately 0.5 at any time prior to the third dose. The ability to stratify patients at these early stages, especially before the first dose of drug, is invaluable for clinicians to make timely and informed decisions regarding course of treatment. This result suggests that our model detects a response signal that is not the result of drug-induced microbiome perturbations, but rather belongs to the persistent individual microbiome signature.

To provide additional evidence in support of detecting individualized response signal in microbiomes by our predictor, independently of drug-induced microbiome-restructuring effects, we provide per-sample visualization of prediction model calls (Figure 5C). These per-sample prediction visualizations, stacked by subject on top of ground truth response indication, allow for analysis of per-subject dynamics in response prediction signals.

It is easily noticeable that in the absolute majority of cases, the sign of response prediction, despite changes in confidence level, remained the same, with one prominent exception: subject #6 of the test set. This suggests that our model captures properties of individual microbiomes per se, as opposed to responding to drug-induced microbiota perturbations as seen in the original publication. Simultaneously, our model captures the latter as part of the overall signal, evident in the evolution of confidence calls following increasing number of drug doses, as observed in patients 24 and 31 (training set) and 6 and 13 (test set). This phenomenon contributes to the overall increase in classification performance after the 3rd dose (Figure 5B), resulting in correct prediction of responder status for the majority of patients based on their last fecal sample (87.5% and 72.7% on training and test sets, respectively).

The impressive overall stability of R/NR binary calls by the model for individual subjects captures individualized clinically relevant microbiome signatures as the primary part of the detected signal. To add an intuitive numerical insight, subjects who received the longest treatment followed by microbiome samples (14 doses in training set subject #14 or 12 doses in test set subject #25) had single-direction response prediction in all samples belonging to a subject, despite having a potential of 2^14^ = 16,384 or 2^12^ = 4,096 combinations of binary classes if classification were performed randomly.

### Lookup-table-based standalone prediction engine

To translate the predictive model into a deployment-ready clinical tool, we distilled the neural network into a compact lookup table (LUT) optimized to preserve predictive power while minimizing computational requirements. This approach decouples the prediction engine from the neural network infrastructure, enabling standalone deployment in clinical settings without specialized hardware or software dependencies.

The LUT was constructed through frugal adaptive quantization of the normalized abundance values for each of the eight features (Table 2). The number of quantization levels per feature ranged from 2 to 3, reflecting the informativeness distribution of each biomarker’s abundance range. This frugal quantization resulted in a compact table of 2,916 key combinations (the product of per-feature levels: 2 × 3 × 3 × 2 × 3 × 3 × 3 × 3 = 2,916), each associated with a predicted therapy response probability.

**Table 2.** Frugal adaptive quantization of normalized abundance values for each model feature. Levels indicate the number of quantization breakpoints; key abundance values define the LUT grid vertices.

| Feature | Levels | Key abundance values |
| --- | --- | --- |
| *Acidaminococcaceae* | 2 | 0.0, 1.0 |
| *Ligilactobacillus* | 3 | 0.0, 0.76, 1.0 |
| *Pasteurellaceae* | 3 | 0.0, 0.7, 1.0 |
| *Dorea* | 2 | 0.0, 1.0 |
| *Barnesiella* | 3 | 0.0, 0.1, 1.0 |
| *Tannerellaceae* | 3 | 0.0, 0.56, 1.0 |
| *Ruminococcus* | 3 | 0.0, 0.33, 1.0 |
| *Atopobium* | 3 | 0.0, 0.79, 1.0 |

To ensure reliable performance on unseen datasets, the LUT-based predictor employs a frozen normalization pipeline. Input data is first subjected to a Centered Log-Ratio (CLR) transform computed over all taxa in the sample, followed by extraction of the 8 model features and min-max scaling using frozen boundaries derived from the discovery cohort (Table 1). This frozen pipeline guarantees that normalized values are not biased by the structure of the particular dataset being analyzed, a critical requirement for clinical deployment where batch effects and protocol variations are common.

Interpolation between LUT vertices leverages N-linear interpolation [31] performed in unbounded space rather than directly in the bounded [0, 1] probability space, improving accuracy in the inherently non-linear probability domain [27]. The interpolated log-probability values are mapped back to valid probabilities via the Softmax transformation [28].

The LUT-based predictor faithfully reproduced the discrimination performance of the original neural network. ROC curve analysis (using a single representative model from the 100-model distribution overviewed in Figure 5A) demonstrated close correspondence between the neural network and LUT predictions (Figure 7A). Scatter plots of LUT-predicted versus neural network-predicted response probabilities confirmed strong agreement, with predictions colored by ground-truth therapy response showing clear separation between R and NR groups (Figure 7B). That end-to-end pipeline test, from separate unnormalized tables of genus and family-level counts through the frozen CLR normalization, min-max scaling, LUT lookup, unbounded space interpolation, confirmed that the complete standalone system produces therapy response probabilities consistent with the original model, with model-to-LUT prediction correspondence observed across the entire dataset (Figure 7C).

**Figure 7:**
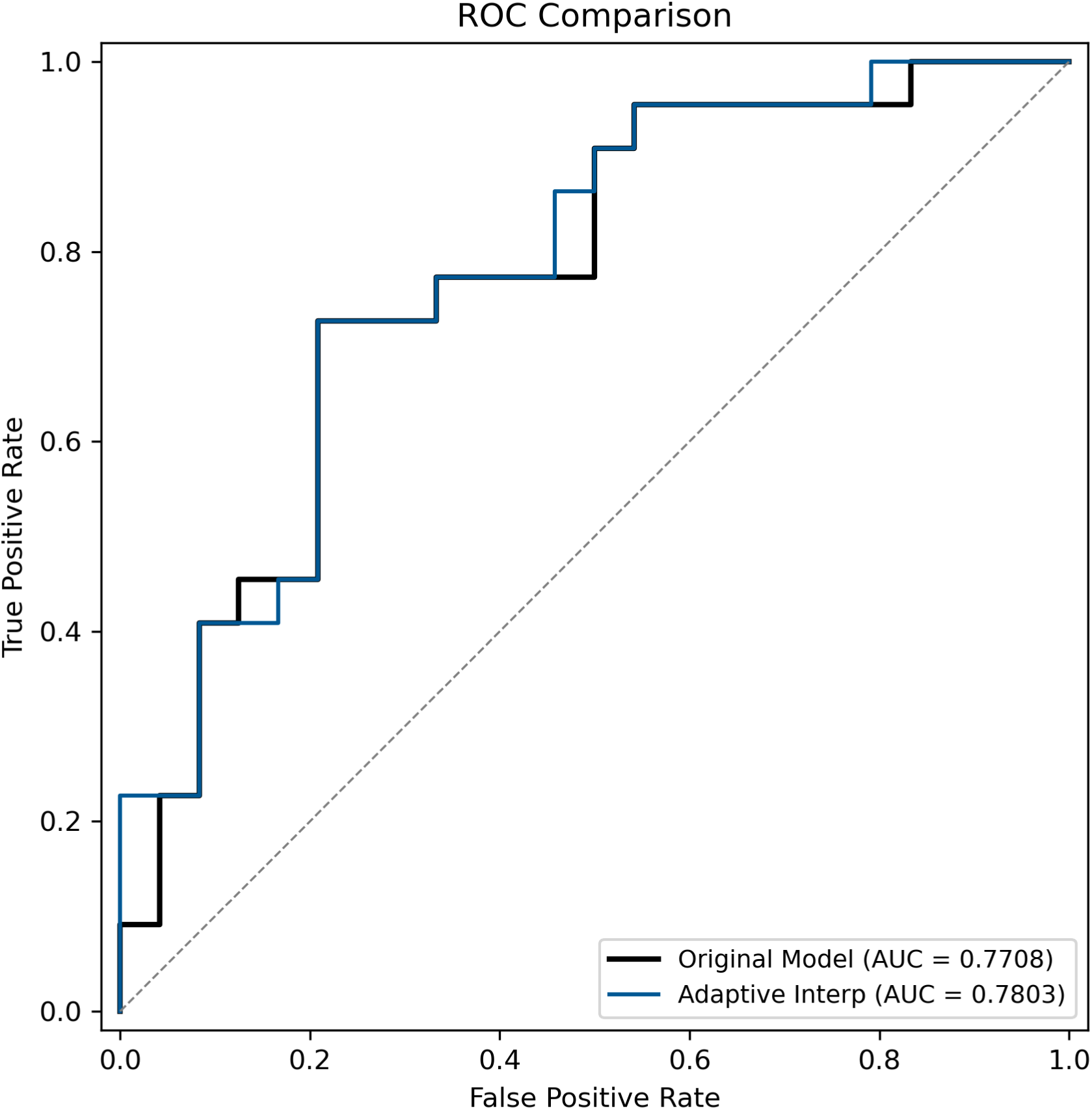
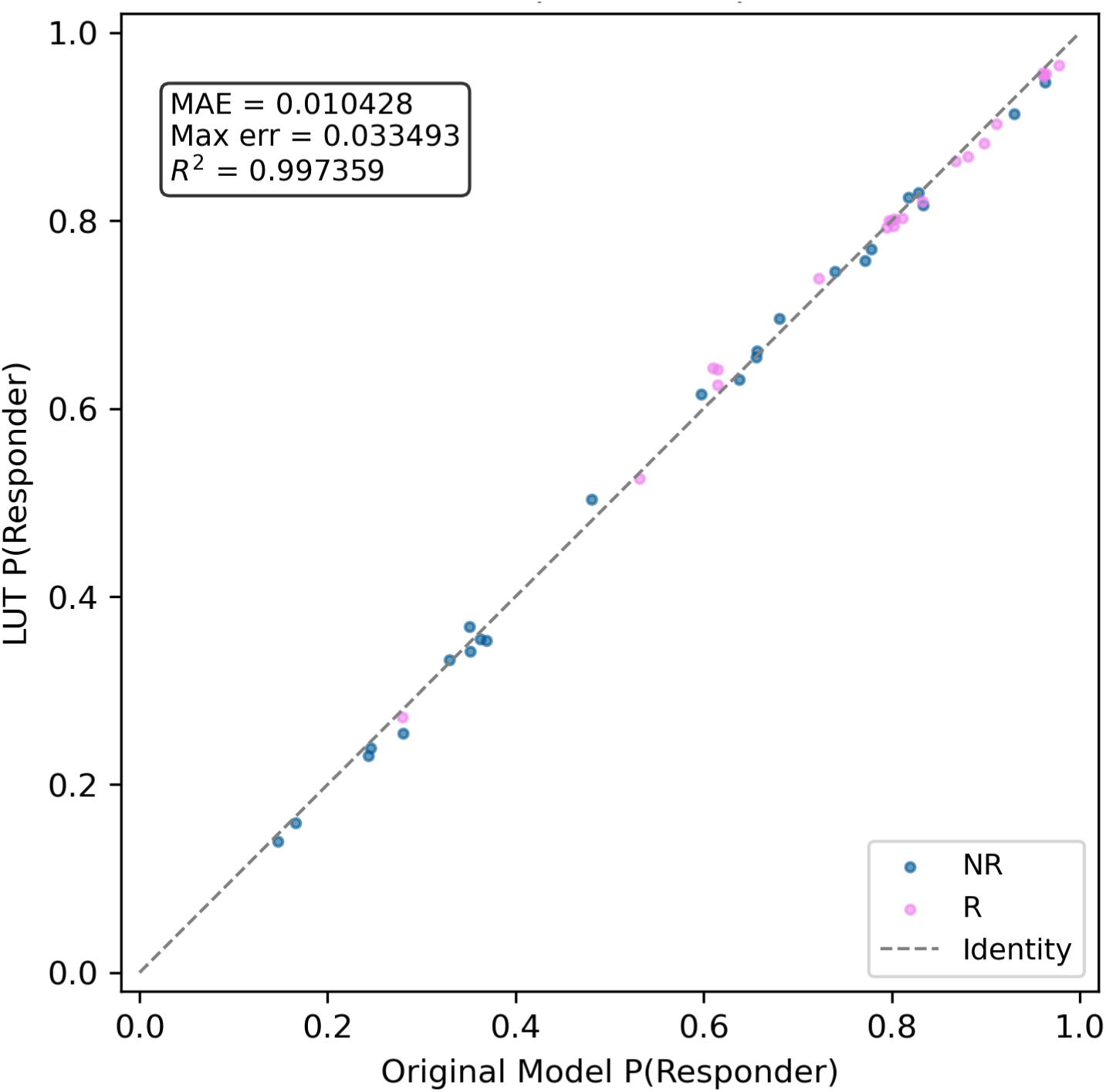
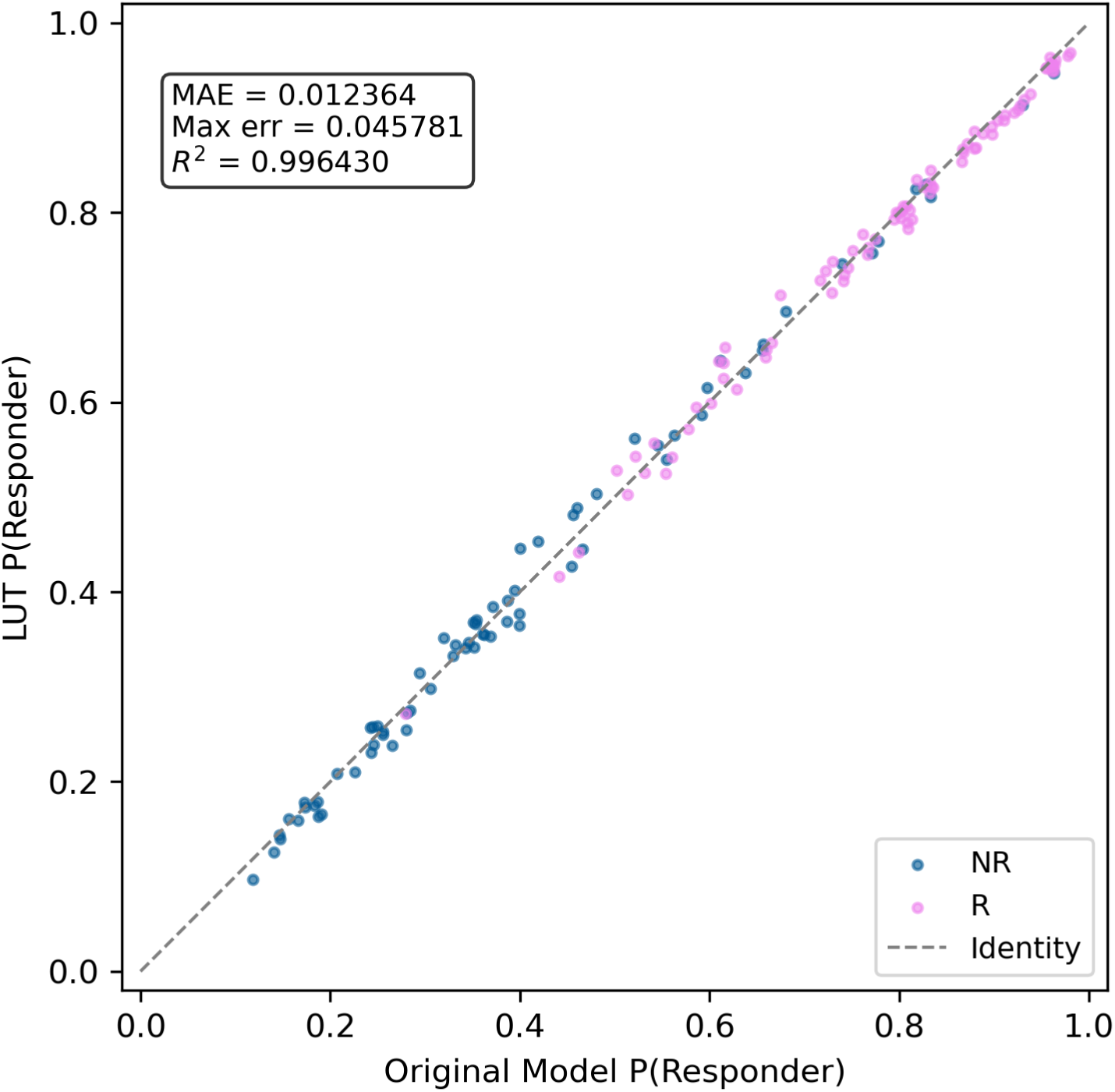
LUT-based prediction engine performance. (A) ROC curves comparing the original neural network model (black) and LUT-based predictions (blue) in the test data split. (B) Correspondence between LUT-based prediction values and the values output by the original model in the test data split. (C) Correspondence between LUT-based prediction values and the values output by the original model in the entire dataset. Dots are colored by reported therapy response.

The LUT-based deployment approach offers several advantages for clinical translation: (1) it eliminates the need for neural network inference infrastructure, (2) the frozen normalization constants ensure reproducibility across clinical sites and analysis centers, (3) the compact table size (2,916 rows) enables implementation in resource-constrained environments, and (4) the explicit representation of all input-output mappings facilitates regulatory review, interpretability and clinical validation.

### Limitations

Several limitations should be acknowledged. First, this study represents an analysis of a single cohort of 35 patients, and external validation in independent cohorts is essential before clinical implementation. The baseline AUROC of 0.681, while significantly above random chance and a qualitative advance over prior work, requires improvement for robust clinical decision-making. Second, the neural network architecture, while optimized for this dataset, may require recalibration for different patient populations or sequencing protocols. Third, the proprietary nature of portions of our feature selection pipeline limits full reproducibility, though the identified 8-taxon signature, the LUT with its frozen normalization constants, and the general methodological framework are fully disclosed. Fourth, the cohort was predominantly male (89%) and of a single geographic origin, potentially limiting generalizability.

## Conclusion

We have shown that, despite the well-established, notoriously large inter-subject variation in gut microbiome composition, it is possible to create a performant drug response predictor (that balances informativeness and prevalence of individual features) by selecting a representative mix of features across taxonomic ranks and using them as inputs to a custom-optimized neural network. The resulting model is advantageous compared to other models reported in the area because of a combination of (1) its capability to stratify subjects before drug intervention, relying largely on innate composition of individual microbiome rather than drug-induced microbiome perturbations, and (2) doing so in a parsimonious manner, without requiring a long list of biomarkers or expensive whole genome, strain-level data resolution. The distillation of the trained model into a portable lookup table with frozen normalization parameters provides a concrete framework for prospective validation and standalone clinical deployment.

Translation of these findings will require: (1) prospective validation in geographically diverse cohorts, (2) development of a standardized assay protocol for the 8-taxon signature, (3) integration with clinical and molecular predictors in multimodal frameworks, and (4) clinical evaluation of the LUT-based prediction pipeline in a point-of-care setting. The parsimonious nature of the signature (8 taxa expected to be detectable by streamlined approaches such as a targeted qPCR) and the self-contained nature of the LUT-based predictor facilitate development of a practical clinical test that can be deployed independently of the computational infrastructure used for model discovery.

## Data Availability

The data that support the findings of this study (deposited by the authors of the original study) are openly available in NCBI Sequence Read Archive, BioProject PRJCA006814.

https://ngdc.cncb.ac.cn/bioproject/browse/PRJCA006814

## Supplements

**Supplementary Table 1: the LUT.** 2,916 rows of the LUT are generated by exhaustive combination of each of the 8 biomarker abundance values at every quantization breakpoint (summarized in Table 2), ready for an adaptive interpolation procedure to handle any real-world combination of biomarker abundance values.

**Supplementary Figure 1: Number of Anti-PD-1 doses received by each patient.**

Barplot showing the number of anti-PD-1 doses received by each patient. Pink bars represent responders (R), and blue bars represent non-responder (NR). The plot highlights the variability in the number of doses administered to each patient. The identifiers of the patients on the x-axis are arbitrary and based on the order of samples given in the metadata.

**Supplementary Figure 2: Shannon diversity analysis of microbial communities.**

Boxplots showing Shannon diversity indices for microbial communities at different taxonomic levels (phylum, class, order, family, genus) for responder (R) and non-responder (NR). The upper row represents samples taken at baseline, while the lower row includes all samples. Pink boxes represent responders, and blue boxes represent non-responders. No significant differences in Shannon diversity were observed between R and NR at any taxonomic level.

**Supplementary Figure 3: Principal Coordinate Analysis (PCoA) of microbial community composition.**

Principal Coordinate Analysis (PCoA) plots depicting beta diversity measures using Bray-Curtis dissimilarity, Jaccard distance, and weighted UniFrac for responders (R) and non-responders (NR). Pink dots represent R and blue dots represent NR. The upper row shows the PCoA plots at baseline. The principal coordinates, PCo1 and PCo2 on the x- and y-axis, respectively, represent the top two axes of variance in the microbial community composition. The lower row shows the PCoA plots for all samples.

**Supplementary Figure 4: Baseline microbiota composition of the study subjects**

Stacked bar plots showing the proportions of microbiota at four taxonomic levels - phylum, class, order, and family - before anti-PD-1 intervention in non-responders (NR) and responders (R). For all plots, each bar represents an individual patient, with colors indicating different taxa. Only the top 8 taxa by overall abundance are annotated, with aggregation of the lower-abundant taxa as “Others”.

## Financial support and sponsorship

none

## Conflicts of interest

The authors are either employees or shareholders of BluMaiden Biosciences. The authors declare no additional competing interests.

## Abbreviations

AFP: alpha-fetoprotein
AUROC: Area Under the Receiver Operating Characteristic curve
CLR: Centered Log-Ratio
CR: Complete Response
GSA: Genome Sequence Archive
HCC: Hepatocellular Carcinoma
ICI: immune checkpoint inhibitors
LUT: Lookup Table
ML: Machine Learning
NGDC: National Genomics Data Center
NR: Non-Responder group
PCoA: Principal Coordinate Analysis
PERMANOVA: Permutational Multivariate Analysis of Variance
PD-1: Programmed Cell Death Protein 1
PR: Partial Response
R: Responder group
SD: Stable Disease
SHAP: Shapley Additive exPlanations
TPOT: Tree-based Pipeline Optimization Tool
TMB: Tumor Mutational Burden
UniFrac: Unique Fraction metric

## Data availability statement

The data that support the findings of this study (deposited by the authors of the original study [1]) are openly available in NCBI Sequence Read Archive, BioProject PRJCA006814.

## Code availability statement

The BluMaiden KEYSTONE™ pipeline is proprietary software developed by BluMaiden Biosciences; its source code is not publicly available, as it is protected intellectual property. The foundational KEYSTONE™ method is the subject of a pending Singapore patent application (No. 10202600190R), and its application to immune checkpoint inhibitor response prediction, as implemented in this work, is covered by a pending international patent application filed under the Patent Cooperation Treaty (PCT/EP2026/054956).

## Author contributions

PCC has designed and executed analysis, interpreted results and drafted the manuscript. OVM designed and supervised the study, interpreted the results, performed the LUT development and wrote the manuscript. RW advised on study design and microbiome analysis procedures and edited the manuscript. DK designed and supervised the project, interpreted the results and edited the manuscript.

## Competing Interests

All the authors are either employees or shareholders of BluMaiden Biosciences. All the authors are inventors on patent applications (Application No. 10202600190R and PCT/EP2026/054956) held by BluMaiden Biosciences Pte. Ltd., which covers the methods, biomarkers and clinical applications described in this manuscript. The authors declare no additional competing interests.

## Notes

### Competing Interest Statement

All the authors are either current or former employees or shareholders of BluMaiden Biosciences. The authors declare no additional competing interests.

### Funding Statement

This study did not receive any funding

### Author Declarations

The study used (or will use) ONLY openly available human data that were originally deposited to NCBI Sequence Read Archive, BioProject PRJCA006814

### Summary of Updates

An extra section was added: delivery of the model's predictions via a Lookup Table. This simplifies the clinical application (and provides an extra layer of transparency and explainability) without the need to deploy the original neural network-based model. The title was also updated accordingly. Additional relevant illustrations proving the LUT applicability were added and discussed in the text.

