## Supplementary figures and images for "Early Prediction of Anti-PD-1 Response in Hepatocellular Carcinoma via Multi-Rank Taxonomic Feature Engineering and a LUT-Based Prediction Vehicle"

### Supplementary Figure 1

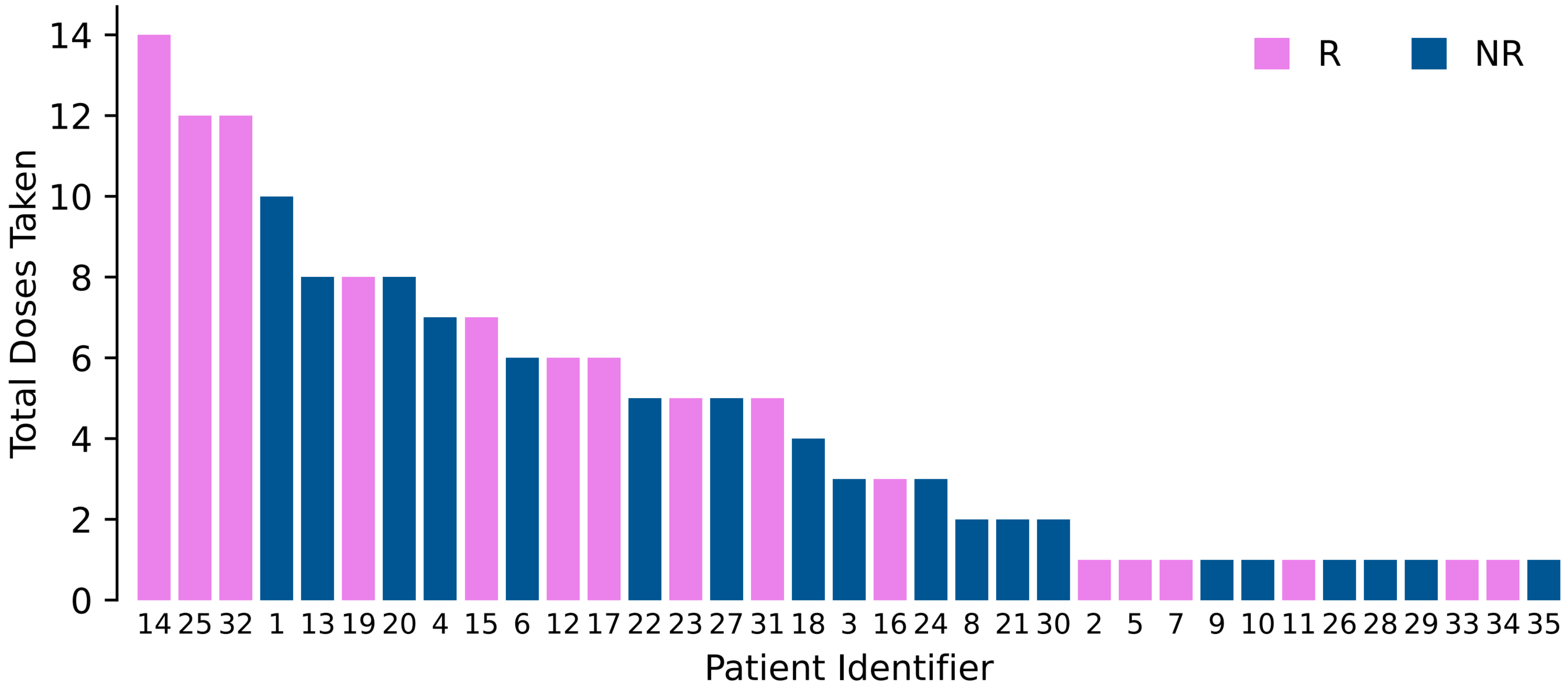

### Supplementary Figure 2

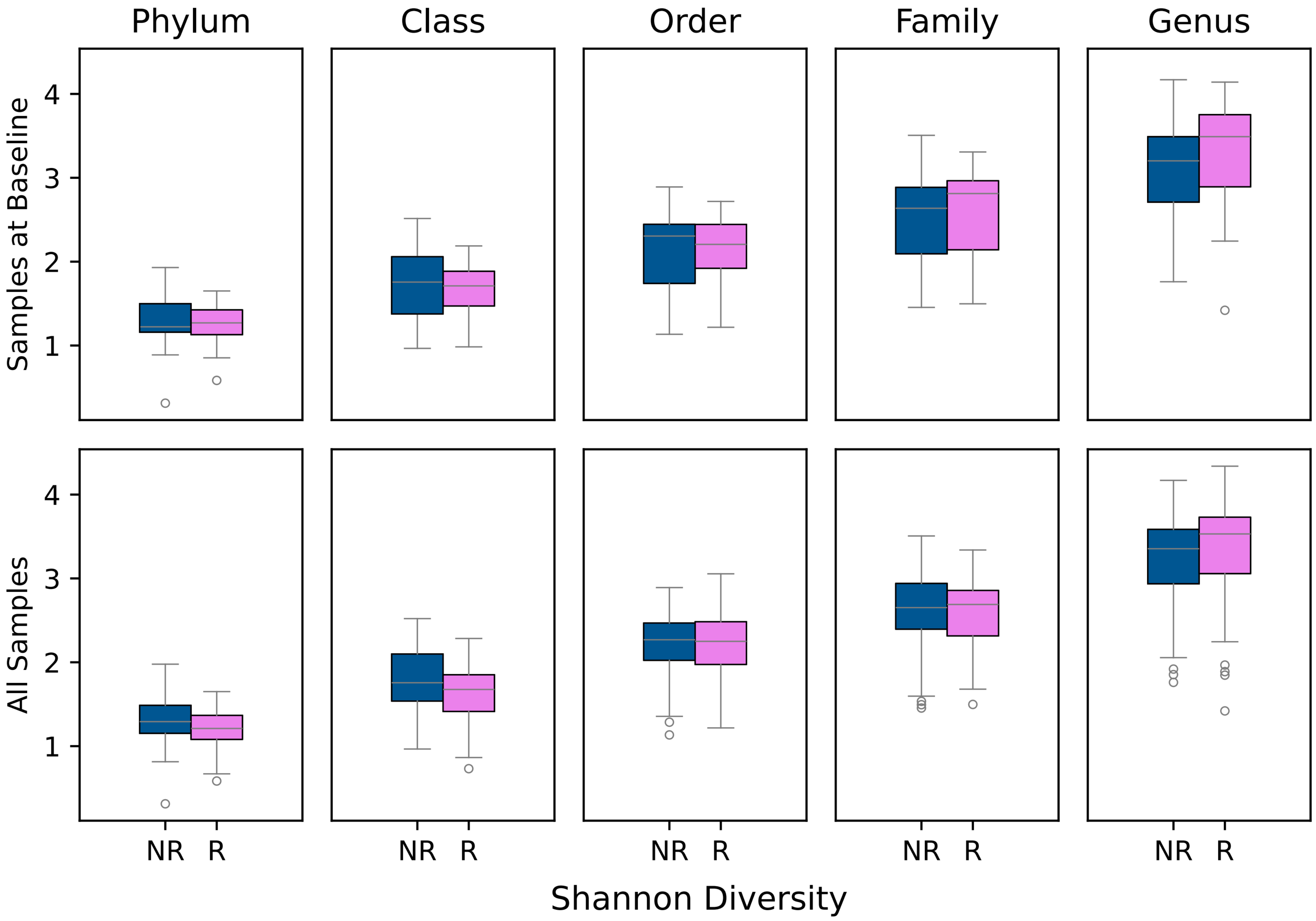

### Supplementary Figure 3

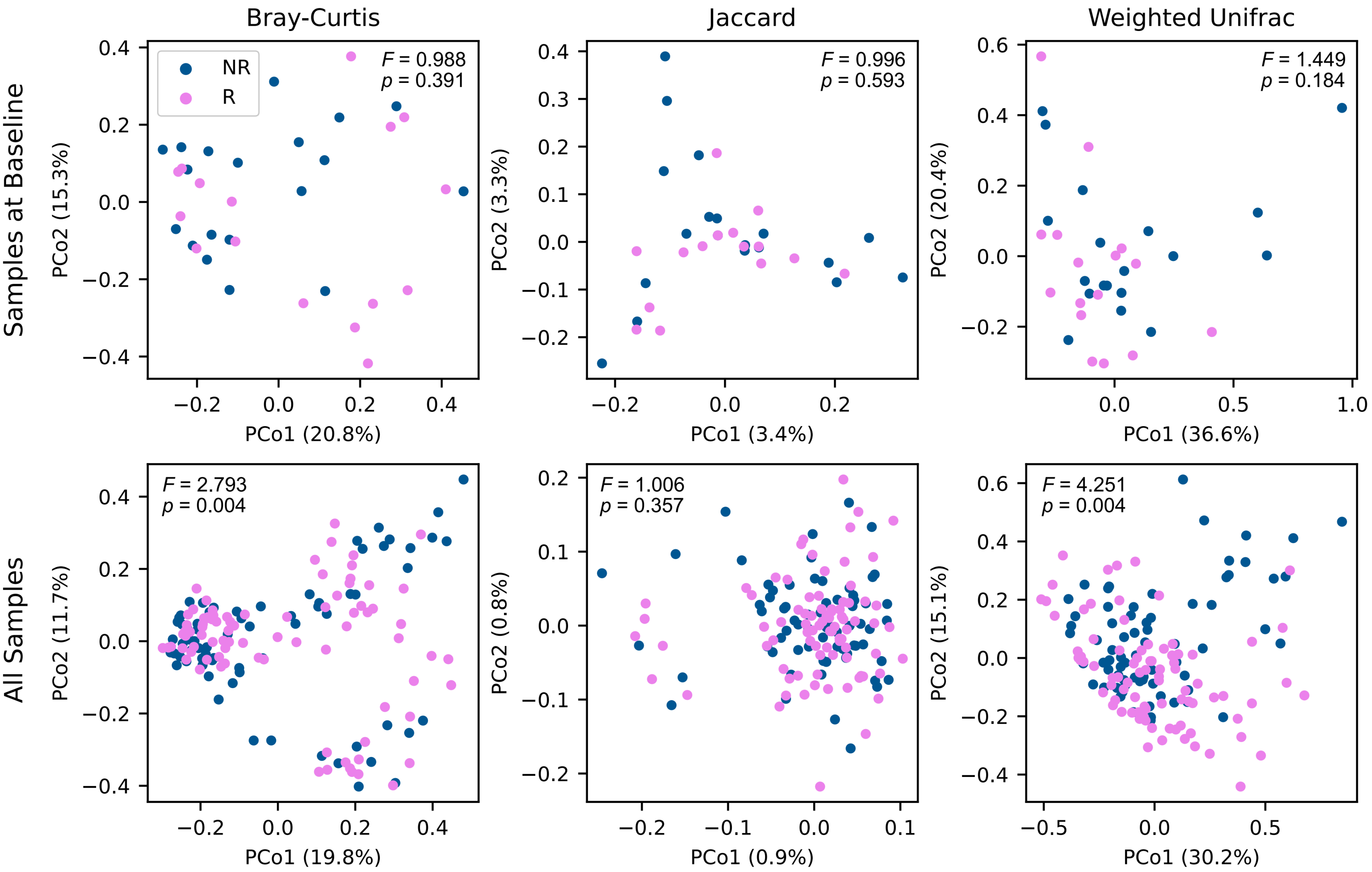

### Supplementary Figure 4

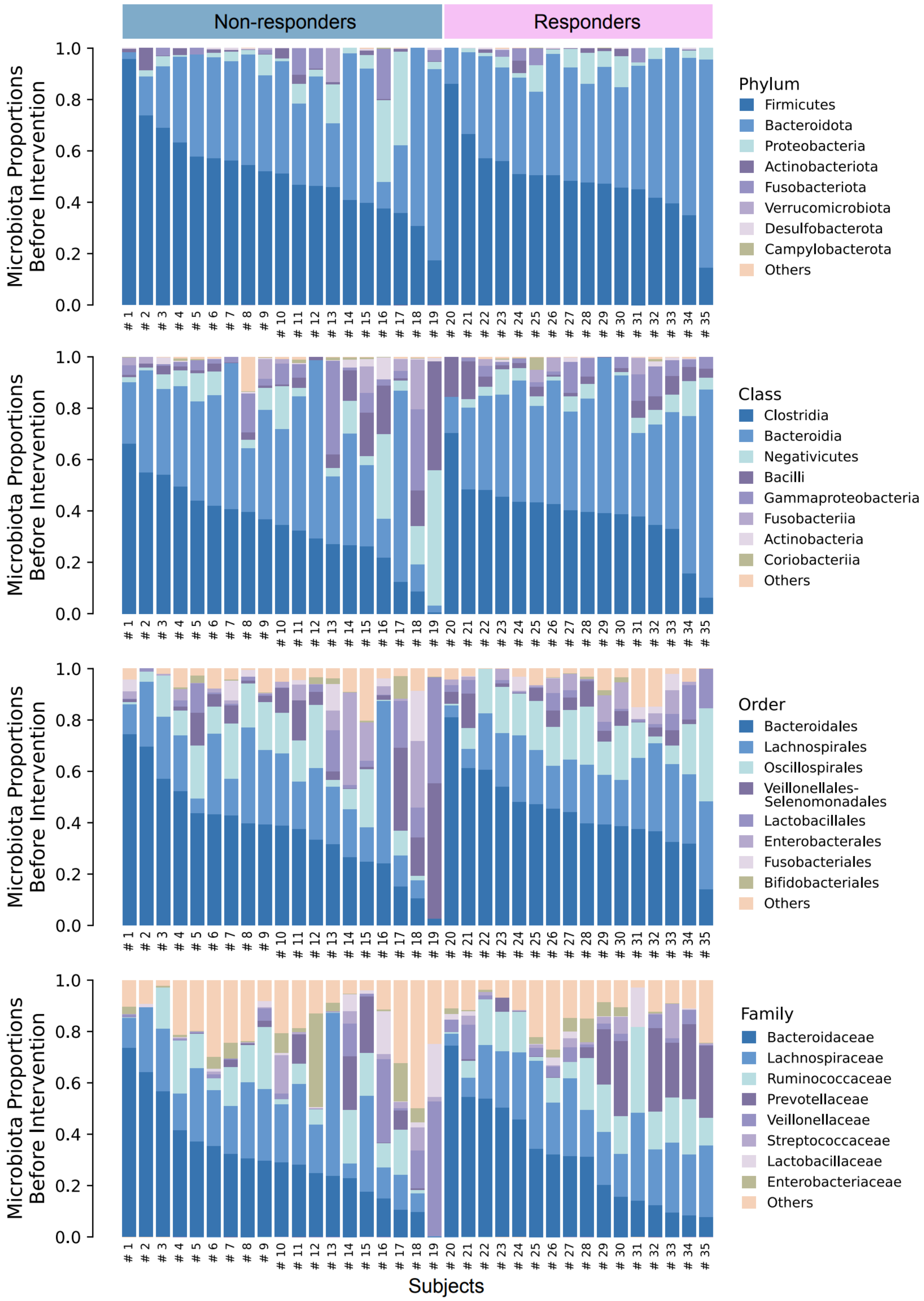
